# An agent-based epidemic model REINA for COVID-19 to identify destructive policies

**DOI:** 10.1101/2020.04.09.20047498

**Authors:** Jouni T. Tuomisto, Juha Yrjölä, Mikko Kolehmainen, Juhani Bonsdorff, Jami Pekkanen, Tero Tikkanen

**Affiliations:** Kausal Ltd, Helsinki, Finland; Finnish Institute for Health and Welfare, Kuopio, Finland; University of Eastern Finland, Kuopio, Finland; Stretta Capital Ltd, Helsinki, Finland; University of Helsinki, Helsinki, Finland

## Abstract

**Background:** Countries have adopted disparate policies in tackling the COVID-19 coronavirus pandemic. For example, South Korea started a vigorous campaign to suppress the virus by testing patients with respiratory symptoms and tracing and isolating all their contacts, and many European countries are trying to slow down the spread of the virus with varying degrees of shutdowns. There is clearly a need for a model that can realistically simulate different policy actions and their impacts on the disease and health care capacity in a country or a region. Specifically, there is a need to identify *destructive policies*, i.e. policies that are, based on scientific knowledge, worse than an alternative and should not be implemented.

**Methods:** We developed an agent-based model (REINA) using Python and accelerated it by the Cython optimising static compiler. It follows a population over time at individual level at different stages of the disease and estimates the number of patients in hospitals and in intensive care. It estimates death rates and counts based on the treatment available. Any number of interventions can be added on the timeline from a selection including e.g. physical isolation, testing and tracing, and controlling the amount of cases entering the area. The model has open source code and runs online.

**Results:** The model uses the demographics of the Helsinki University Hospital region (1.6 million inhabitants). A *mitigation strategy* aims to slow down the spread of the epidemic to maintain the hospital capacity by implementing mobility restrictions. A *suppression strategy* initially consists of the same restrictions but also aggressive testing, tracing, and isolating all coronavirus positive patients and their contacts. The modelling starting point is 2020-02-18. The strategies follow the actual situation until 2020-04-06 and then diverge. The default mitigation scenario with variable 30–40% mobility reduction appears to delay the peak of the epidemic (as intended) but not suppress the disease. In the suppression strategy, active testing and tracing of patients with symptoms and their contacts is implemented in addition to 20–25% mobility reduction. This results in a reduction of the cumulative number of infected individuals from 820 000 to 80 000 and the number of deaths from 6000 to only 640, when compared with the mitigation strategy (during the first year of the epidemic).

**Discussion:** The agent-based model (REINA) can be used to simulate epidemic outcomes for various types of policy actions on a timeline. Our results lend support to the strategy of *combining comprehensive testing, contact tracing and targeted isolation measures* with social isolation measures. While social isolation is important in the early stages to prevent explosive growth, relying on social isolation alone (the mitigation strategy) appears to be a destructive policy. The open-source nature of the model facilitates rapid further development. The flexibility of the modelling logic supports the future implementation of several already identified refinements in terms of more realistic population models and new types of more specific policy interventions. Improving estimates of epidemic parameters will make it possible to improve modelling accuracy further.

## Background

Countries in Asia, Europe, and the Americas have taken different approaches to tackling the COVID-19 pandemic (caused by the novel coronavirus labelled SARS-CoV-2). There are disparate views about whether the pandemic is stoppable and which crucial actions should be taken. For example, South Korea has done extensive testing of symptomatic people and their contacts, including asymptomatic ones, and then isolated everyone who had a positive test result (Normile, 2020). In contrast, Great Britain was originally going for minimal action policy with an idea to let people develop herd immunity, but only a week later they switched to strict social isolation policy (Boseley, 2020).

There are two main approaches to the coronavirus crisis. First, one can assume that a pandemic is already so widespread globally that it will inevitably infect a large fraction of the population. Then the key thing is to slow down the spread in each country so that the health care system can cope with the COVID-19 patients, many of which need intensive care with respirators. Eventually, the epidemic fades away due to herd immunity, i.e. there are enough recovered people to prevent effective spread of the virus in the population. This is called the **mitigation** strategy and often referred to as “flattening the curve”. Depending on the still debated fatality and intensive care rates of COVID-19, the mitigation strategy may require a very long time frame of slow-down measures to allow the health care system to cope. This in turn could lead to unacceptable damage to the economy.

The other main approach assumes that the disease can (and must) be suppressed so that only a small number of patients shall exist in the population. Rather than relying on herd immunity like in mitigation, suppression has to be actively maintained by isolating infected people. *Total* eradication in any one country is impossible in practice as long as the disease is still common in other countries. The epidemic is kept under control by actively searching for all patients (including those without symptoms) and isolating them so that the infection chains are cut. This is called the **suppression** strategy, sometimes referred to as “the hammer and the dance” in mass media. The “hammer” refers to concerted initial actions, such as mobility restrictions and coronavirus testing to get the numbers of infected patients down, and the “dance” is a delicate balance of opening up the society and partially removing mobility restrictions while still maintaining the capacity to trace and isolate all patients (Pueyo, 2020). The dance may continue for months or even years, until a treatment or vaccination has been developed.

The World Health Organisation (WHO) urges countries to *suppress* the disease and get the numbers of active patients down to hundreds. Therefore, WHO strongly recommends that countries increase their coronavirus testing capacity quickly, test their populations aggressively, and trace chains of infection to systematically isolate the infectious individuals (Ghebreyesus, 2020).

Each of the two strategies may contain identical actions at a particular time point. In the beginning of an epidemic, both strategies could implement social isolation, closing down public spaces, widely testing people, tracing contacts, and isolating all with positive tests. The immediate purpose is the same: to increase social distance and reduce the probability of new infections. However, while a mitigation strategist may be content when the number of new cases stabilises, a suppression strategist would demand more actions to trace asymptomatic individuals to make the numbers plummet. Therefore, it is especially important to analyse a series of actions each strategy would demand.

Scientific understanding of SARS-CoV-2 has increased dramatically in just a few weeks. However, there is still uncertainty about a few key variables that make impact assessment and informed policy difficult. One of these key variables is the fraction of asymptomatic people among all infected. It can have two implications.

The asymptomatic fraction appears to be close to a half, as observed with intensive testing in e.g. the Italian town of Vo’Euganeo and the ship Diamond Princess (Ferguson, 2020; ECDC, 2020). This implies that most infected individuals can be identified and quarantined with active testing of symptomatic patients *and their contacts, since the asymptomatic cases are typically close to symptomatic ones in the chains of infection*.

Despite these data points, it is not rare to read claims that the asymptomatic fraction of COVID-19 may even be very close to 1. This would imply that the disease would be much more widespread in society than confirmed cases, which would reduce population-level consequences due to fewer severe cases, but also make the disease more difficult to suppress. However, with mortality due to COVID-19 reaching already almost 0.1% of the *total population* of Lombardy (Italian Ministry of Health, 2020) it is becoming clear that there is little scope for optimism about the severity of the disease.

The mitigation strategy would be compatible with a very high fraction of asymptomatic cases and the associated reduced severity of the epidemic, while a low-to-moderate asymptomatic fraction both calls for and makes possible a suppression strategy based on comprehensive testing and contact tracing.

This study therefore aims also to provide a timely input into the important Finnish policy discussion. As of writing, Finland’s latest clearly communicated objective is still to just slow down the epidemic, corresponding to the *mitigation* strategy. However, the *suppression* strategy is increasingly discussed by the health authorities and is clearly being evaluated as potentially a better option. Moreover, the very stringent mobility restrictions currently in place appear based on the REINA model to be sufficient to practically eradicate the virus *if continued until fall 2020*. The mitigation strategy would call for relaxation of restrictions, and this has been modelled by under the mitigation strategy simulations discussed in this study.

Tuomisto and coworkers (2020) define “*destructive policies*” as policies that have better alternatives according to scientific knowledge. This concept was used in this work as a criterion to identify whether there were policy strategies that should be abandoned. For example, can it be shown that the suppression strategy (as recommended by WHO) is preferable to the mitigation strategy *regardless* of the currently unknown fraction of asymptomatic infections and other scientific uncertainties?

Typical epidemiological models are based on dividing the population to Susceptible, Exposed, Infectious and Recovered, and modelling the transitions from one category to the other. These models, known as SEIR (or in simpler form SIR) models are typically described (in continuous limit) by ordinary differential equations (e.g. Wilson et al., 2020; Walters et al., 2018). A key parameter in such models is R, the average number of people a patient infects in a population. Its initial value R_0_ describes R in a situation where none of the people have immunity against the disease, so everyone is susceptible.

An agent-based model has a few putative advantages over the traditional models described above. First, there is more flexibility to model certain types of micro-level processes and policy decisions of high practical interest. In particular, modelling of different testing and contact tracing approaches can be done with better precision and ease in agent-based models. In addition, an agent-based model leads naturally to a very straightforward representation of R_0_ (and more generally R_t_) in terms of the infectivity of the pathogen and the interaction characterics of the population. While the former should be largely a property of the pathogen itself, the latter varies across countries. An agent-based model thus lends itself to analysing cross-country variations of epidemic dynamics.

A potential issue with large-scale agent-based models is computational intensity. Indeed one motivation behind this work was to create a proof of concept, demonstrating that agent-based models can be applied to detailed modelling of a large-scale epidemic.

The basic logic of SEIR models can be implemented also as agent-based simulations, which follow agents, or individuals of a population, and their fate through a timeline day by day. The agents interact and get infected, tested, treated, and hospitalized. Policies can be added on the timeline based on actual events or into the future to assess their potential impacts. Additionally, the value of R_t_ (and specifically R_0_) is an emergent property and can be calculated based on the simulation and tracked throughout the epidemic timeline. Such a tool is especially valuable when it is calibrated with an actual population, age structure, contact matrix, and events.

In this work, we developed such an open-source, online model called REINA (Realistic Epidemic Interaction Network Agent model) and adjusted its population-dependent parameters to the Helsinki University Hospital region with 1.6 million people and 1362 confirmed COVID-19 patients at the time of writing (2020-04-05). Our key questions were: Can we identify destructive policies among the two main scenarios or their variants and thus inform policy makers about what should not be done during the weeks to come? What can be learned from the performance and realism of the agent-based model?

## Methods

The model is an agent-based probabilistic simulation model created with Python and accelerated by the Cython optimising static compiler. The time span covers up to two years starting from Feb 18, 2020, with one day intervals. The model uses the population and age structure of the Helsinki University Hospital District (HUS) in Finland. It follows a population over time at individual level during different stages of the disease. It estimates the number of patients in hospitals and in intensive care, including remaining treatment capacity. It also estimates death rates and counts based on the treatment available at each time point.

This model is based on random interactions between agents, and the interactions lead to outcomes with fixed probability distributions (see Table 1). The states are similar to those in SEIR models (e.g. Wilson et al., 2020), which are typically described by differential equations. Although the individual events and times spent in different states are probabilistic, many key parameters whose real-world values still involve significant uncertainty are assumed as known constants in a simulation. Therefore, sensitivity analyses were done with two key parameters: the fraction of asymptomatic infections and the probability of infection given exposure to the virus. In addition, the five ready-made simulations at the web interface were run for 100 times with different random seeds to investigate their sensitivity to random in-model variation.

**Table 1.**
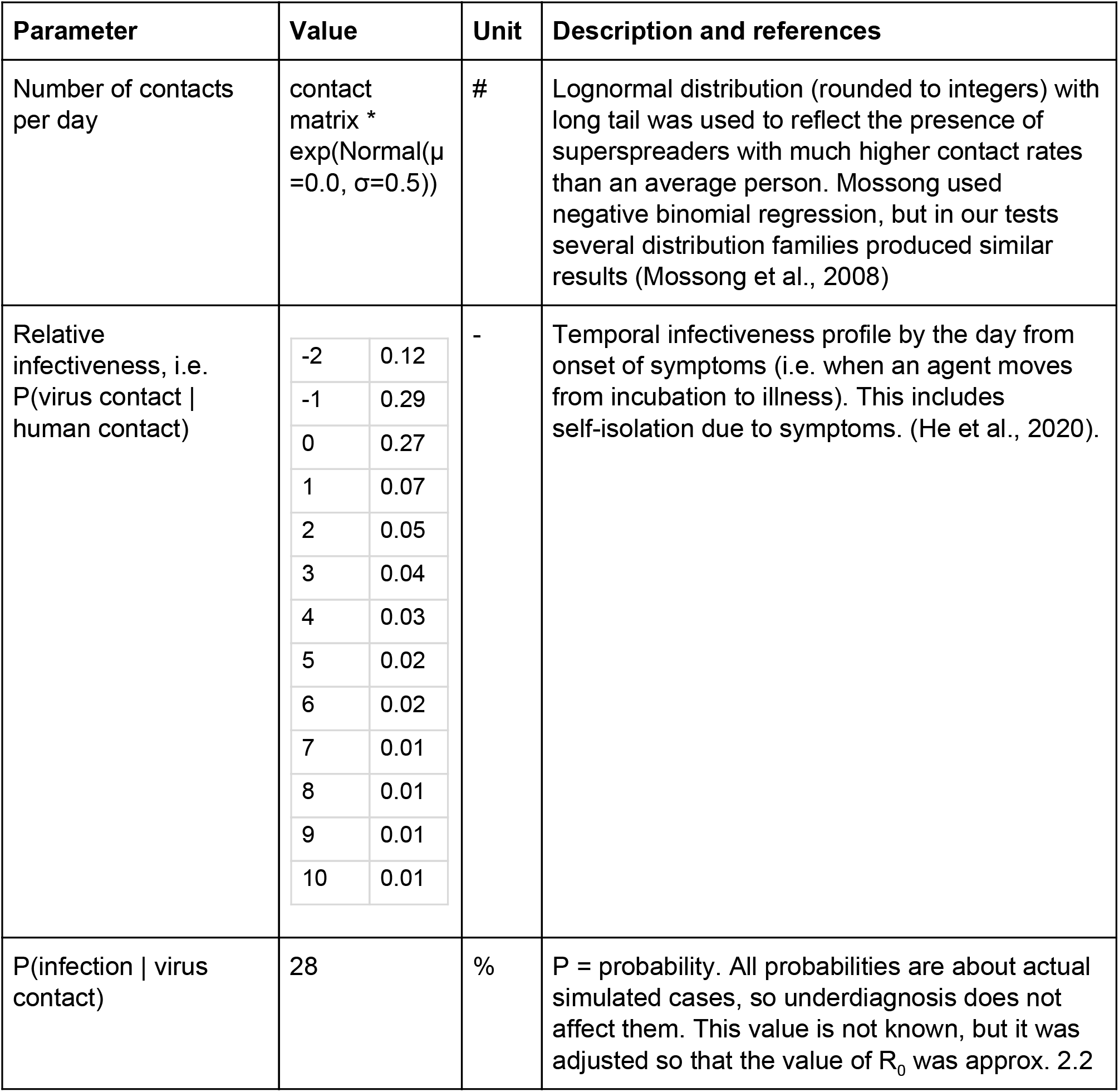
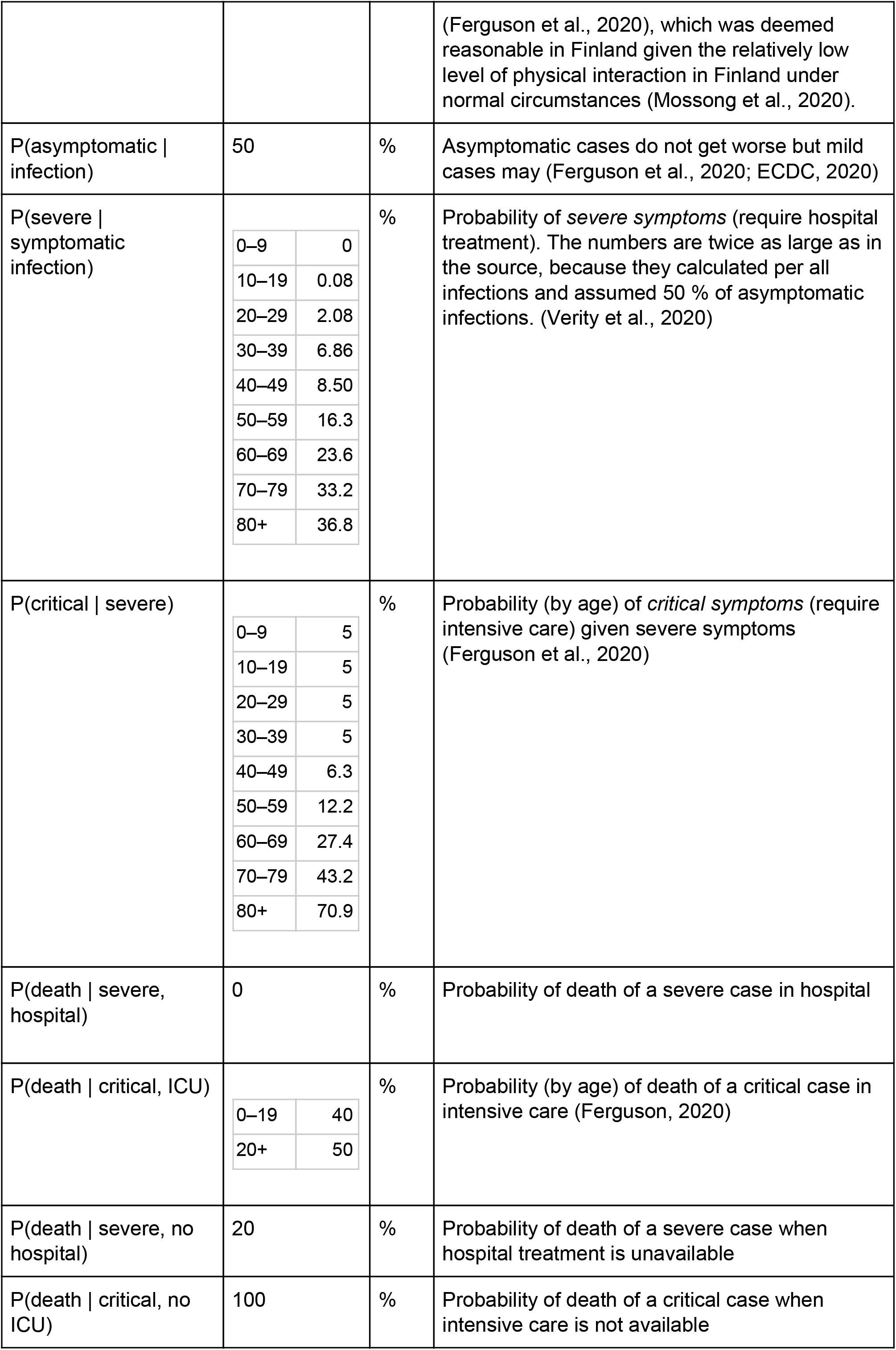
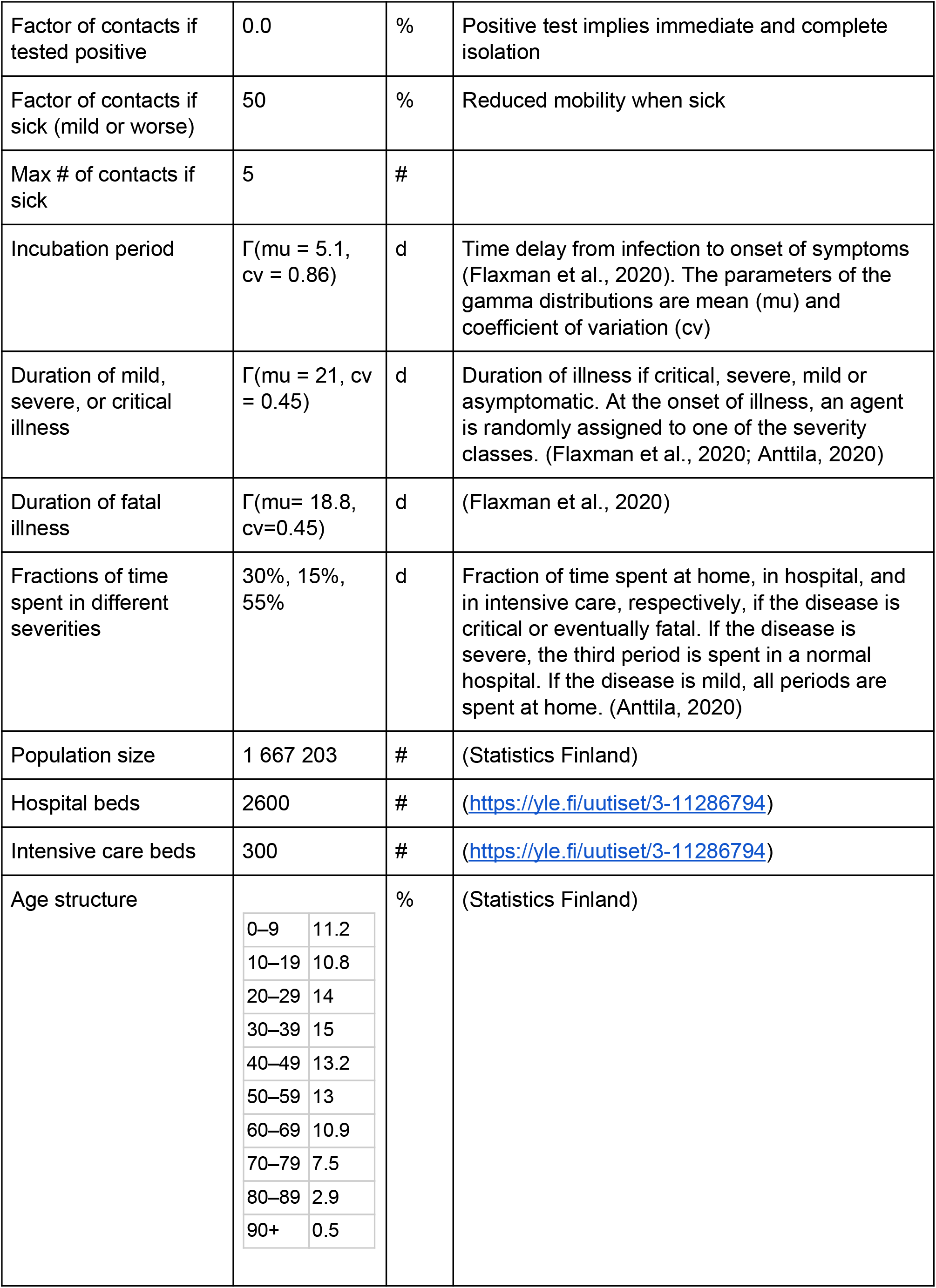
Model parameters for REINA.

**Table 2.** Actions and events available in the REINA model.

| Name | Unit | Description |
| --- | --- | --- |
| Lazy: Test only severe symptoms | - | Coronavirus testing of only those who are hospitalized |
| Active: Test all with symptoms | - | Coronavirus testing of all with respiratory symptoms |
| Aggressive: Test and trace contacts | % | Coronavirus testing of all with respiratory symptoms and, if positive, their contacts. The likelihood of finding an individual contact is given as the parameter. |
| Limit mobility | % | Fraction of mobility (measured as the number of contacts) reduced uniformly for each agent |
| Build new hospital beds | # | Number of hospital beds added to the hospitals |
| Build new ICU units | # | Number of intensive care units added to the hospitals |
| Import infections | # | Number of infected people added to the population (with the same age distribution as existing population) |

**Table 3.** The default scenario used as the basis for all other scenarios. It contains actual decisions and events in Finland from February to March 31st, 2020, and a few projected actions about ICU capacity.

| Date | Event | Description |
| --- | --- | --- |
| 2020-02-20 | Test all with symptoms | Active testing policy |
| 2020-02-22 | Import infections: 5 infections | The numbers of imported infections are estimated based on reported trips from e.g. Italy and Austria and the data of confirmed cases, and their level was estimated by calibrating the beginning of the epidemic timeline. |
| 2020-03-05 | Import infections: 20 infections |  |
| 2020-03-07 | Import infections: 120 infections |  |
| 2020-03-09 | Import infections: 120 infections |  |
| 2020-03-11 | Import infections: 80 infections |  |
| 2020-03-12 | Limit population mobility: 10% | Effectiveness of the actual actions is quantified using author judgement and Google (2020). |
| 2020-03-13 | Import infections: 20 infections |  |
| 2020-03-13 | Test people only with severe symptoms | Lazy testing policy |
| 2020-03-15 | Import infections: 20 infections |  |
| 2020-03-16 |  | Government declares state of emergency |
| 2020-03-17 | Limit population mobility: 20% | Government implements the law of readiness state |
| 2020-03-20 | Limit population mobility: 30% |  |
| 2020-03-22 | Limit population mobility: 35% |  |
| 2020-03-28 | Limit population mobility: 50% | Isolation of the Helsinki region from the rest of the country. Total restriction of all actions based on Google (2020). |

**Table 4.** The two main strategies used in the model: the mitigation strategy to slow down the epidemic, and the suppression strategy recommended by WHO.

| Date | Mitigate<br>(ensure intensive care) | Suppress<br>(Hybrid model) | Description |
| --- | --- | --- | --- |
| 2020-04-06 | Limit mobility: 50% | Limit mobility: 50% | Starting point for the strategies |
| 2020-04-06 | Lazy testing: test severe cases only | Lazy testing: test severe cases only |  |
| 2020-04-30 | Build new ICU units: 150 | Build new ICU units: 150 |  |
| 2020-05-01 |  | **Aggressive testing:**<br>**30% accuracy of**<br>**contact tracing** | Key difference between the strategies |
| 2020-05-01 |  | Limit mobility: 20% | Alleviate restrictions as testing and tracing kicks in |
| 2020-05-30 | Build new ICU units: 150 | Build new ICU units: 150 |  |
| 2020-06-01 |  | Aggressive testing: 40% |  |
| 2020-06-15 | Limit mobility: 30% |  | Adjust restrictions in accordance with strategy goals |
| 2020-06-24 |  | Limit mobility: 25% |  |
| 2020-06-30 | Build new ICU units: 150 |  |  |
| 2020-06-30 | Build new hospital beds: 300 | Aggressive testing: 50% |  |
| 2020-07-15 | Build new ICU units: 150 |  |  |
| 2020-07-15 | Build new hospital beds: 300 |  |  |
| 2020-07-30 | Build new ICU units: 150 |  |  |
| 2020-07-30 | Build new hospital beds: 300 | Aggressive testing: 60% |  |
| 2020-08-15 | Build new ICU units: 150 | Limit mobility: 20% |  |
| 2020-08-15 | Build new hospital beds: 300 |  |  |
| 2020-08-30 | Build new ICU units: 150 |  |  |
| 2020-08-30 | Build new hospital beds: 300 |  |  |
| 2020-09-15 | Limit mobility: 40% |  |  |
| 2020-11-15 | Limit mobility: 30% |  |  |
| 2020-12-15 | Limit mobility: 40% | Limit mobility: 25% |  |

There is a clear user interface where any number of interventions can be added on the timeline from a selection including e.g. mobility restriction, several levels of testing and tracing, and inserting cases into the area. A publicly available instance of the model is online at https://korona.kausal.tech/sim and calculates one scenario in a few seconds. The implementation is open-source and the code is available at https://github.com/kausaltech/reina-model. The release that produced the results of this article is 1.0. The simulation outputs, analysis code, and analysis results and figures of this manuscript are available at https://github.com/jtuomist/corona.

The agents represent individual people, who have the following properties: age (years), infected (yes/no), infection detected (yes/no), immunity (yes/no), days of incubation period left (d), days of illness since the onset of symptoms (d), other people infected (identifiers of agents that this agent infected; used for tracing), state, and symptom severity. The possible states of a person are the following: susceptible, incubation (infected but actual illness not yet started; may spread virus), illness, hospitalized, in intensive care, recovered (i.e., immune), and dead (Figure 1). The symptom severity of a patient can be asymptomatic, mild, severe, or critical. The model follows every individual through the whole disease path from susceptible to recovery or death.

**Figure 1.**
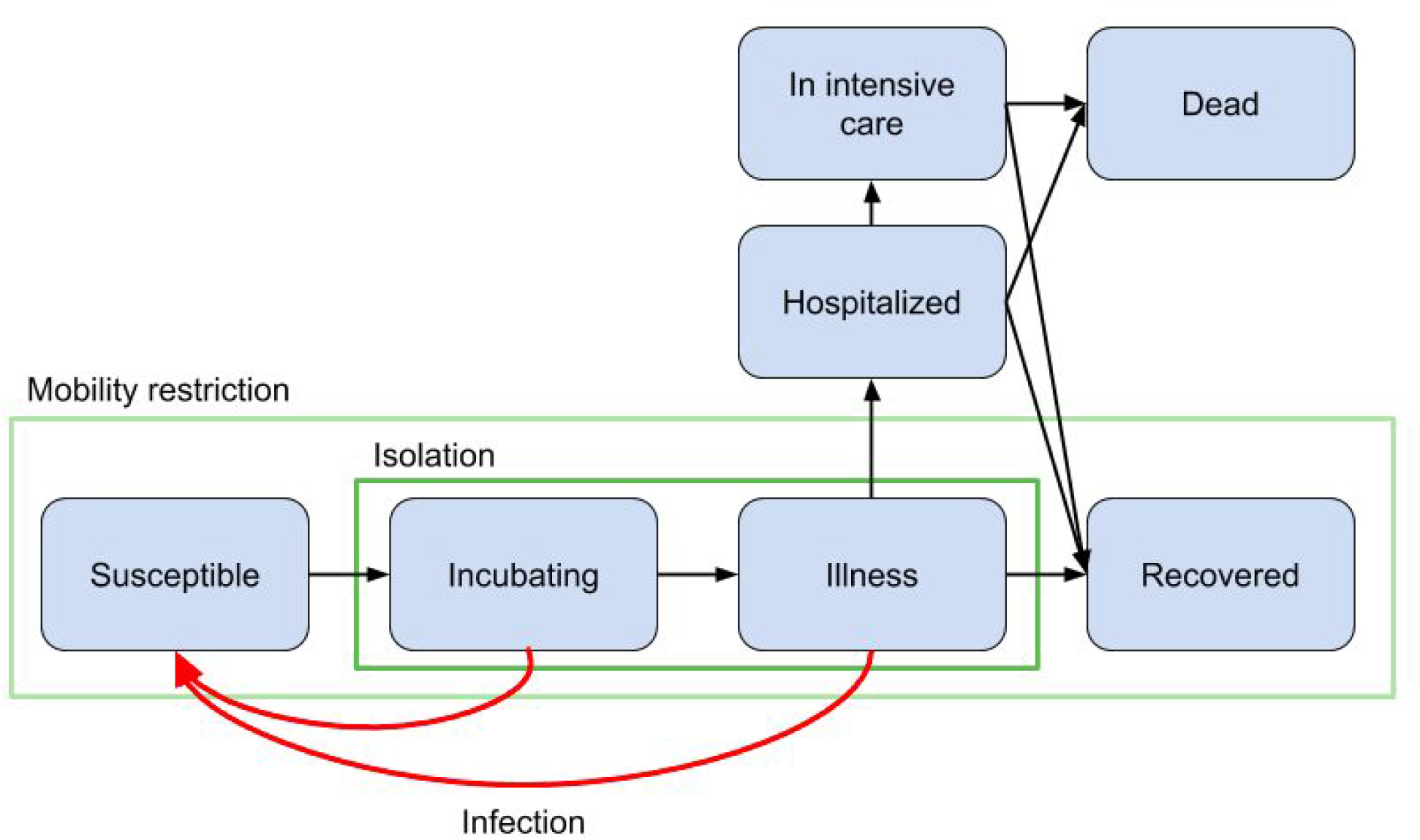
Diagram of the coronavirus agent model. Blue nodes: different states of the agents. Black arrows: transitions between the states. Red arrows: infection pathways. Green boxes: targeted (isolation) and broad (mobility restriction) population subgroups under varying degrees of social isolation. Incubating state contains people that already have the virus and may spread it but the actual illness (i.e., symptoms) has not yet started (see Table 1). In this model, the “exposed” state is treated implicitly, and the “infectious” state is divided into four sub-states. Thus, they are not shown here as state names.

The healthcare system has the following properties: number of hospital beds, number of hospital beds available, number of intensive care beds, number of intensive care beds available, testing mode (lazy: test only people with severe symptoms; active: test also people with mild symptoms; aggressive: test also people with mild symptoms and contacts of coronavirus positive people), number of coronavirus tests run per day, length of testing queue. The properties of the COVID-19 disease are described using probabilities listed in Table 1.

An important part of the model is the contact algorithm. Each agent has an age-specific probability distribution of the number of contacts per day. Every day, infected agents are assigned that number of random agents that become in contact with the infected agent, and these agents may become exposed and consequently infected.

In real life, contacts clearly are not random but clustered by e.g. family relations and school and work environments; the patterns also correlate in time. These important properties are not dealt with in the current version of REINA, as is the case with most epidemic models.

The model keeps track of every contact the infected agents have. This enables the examination of different testing and tracing policies. The current model version can test all agents with symptoms and then perform contact tracing with a given accuracy, which can be adjusted by the user.

The outcome is measured in several ways. The numbers of individuals in each state are followed daily. The numbers of dead and recovered are in practice cumulative because there is no exit from those states. The number of available hospital and intensive care beds are also calculated. The infection fatality rate (IFR) and case fatality rate (CFR) are calculated for each day by dividing the cumulative number of deaths by the cumulative number of total infections or confirmed cases, respectively. The effective reproductive number R_t_ is calculated for each day as the average number of infections caused by agents whose disease ends on that day.

We also developed a new simple indicator for side effects of mobility restriction: *restriction day index* (RDI). It is defined as the cumulative sum of daily percentages of mobility restrictions. For example, a week with 10% restriction and a day with 70% restriction will both produce 0.7 RDI. Although this indicator is far from an optimal measure of societal or economic impacts, it still gives a rough estimate for scenario comparison.

## Results

A general view of the results produced by the REINA model are presented in Figure 2 (mitigation strategy) and Figure 3 (suppression strategy). The two scenarios were created to either slow down the spread of the epidemic or to suppress the number of cases. The scenarios used lazy and aggressive testing policies, respectively, and both scenarios adjusted the mobility restrictions to achieve an acceptable outcome.

**Figure 2.**
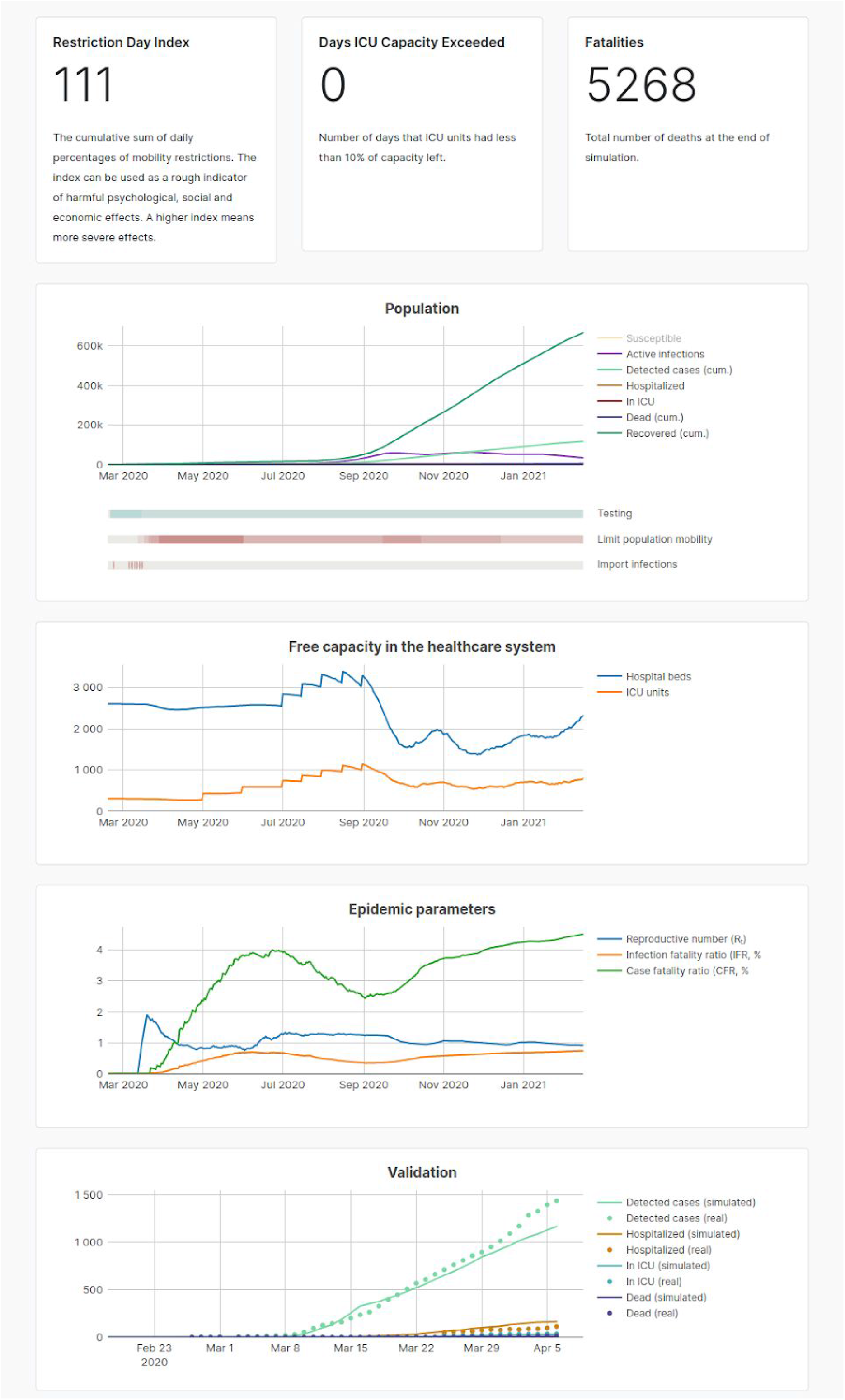
A screenshot from the model output. Timeline of a mitigation strategy (new hospital beds, 30-40% mobility restriction and only severe cases tested). At the end of the one-year simulation, there are 660000 recovered patients (data not shown). The results are for a single simulation run, not an average of an ensemble.

**Figure 3.**
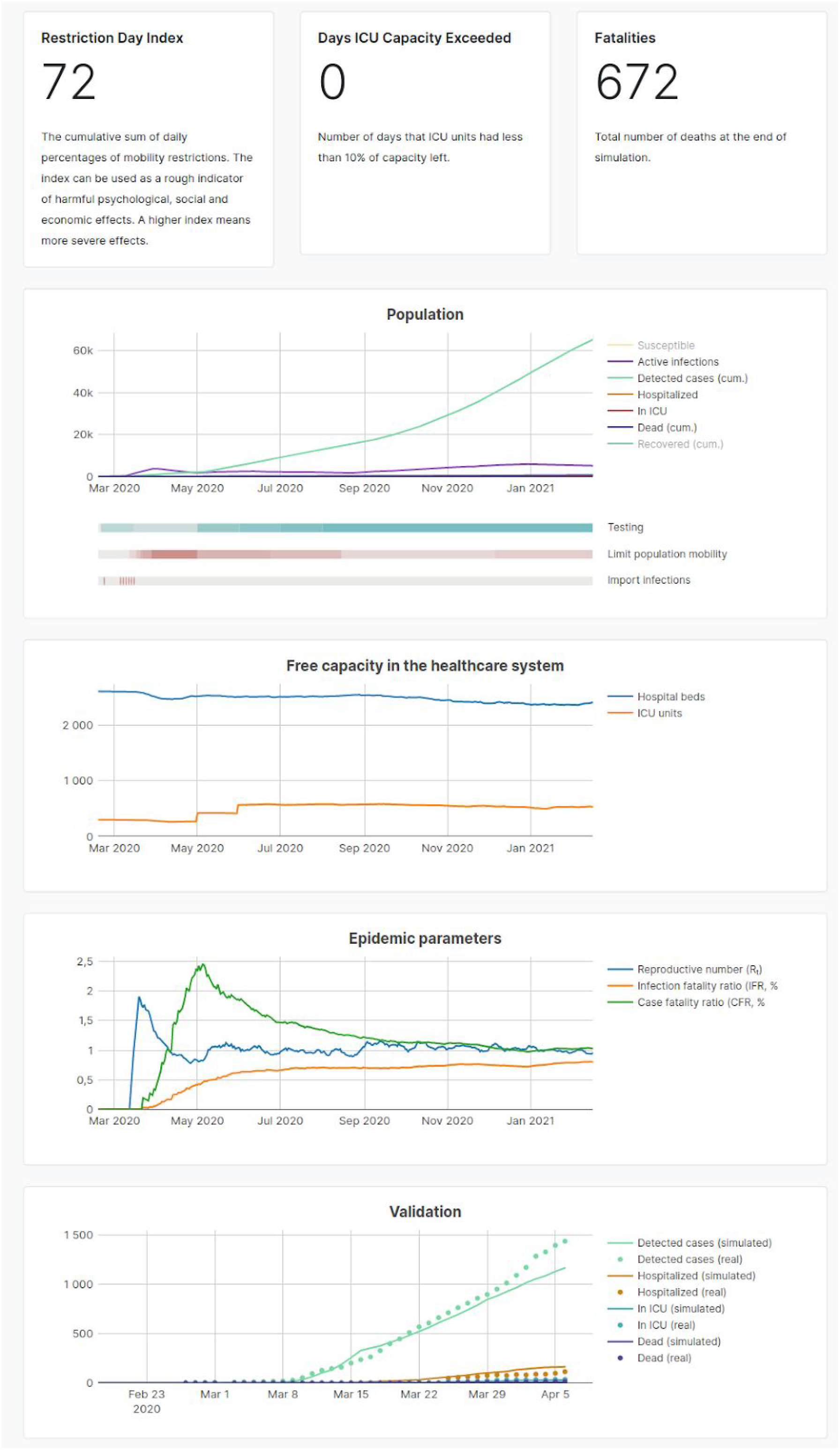
A screenshot from the model output. Timeline of a suppression strategy (initially 50% and later 20-25% mobility reduction with aggressive testing of mild cases and tracing of contacts). At the end of the one-year simulation, there are 77000 recovered patients (data not shown).The results are for a single simulation run, not an average of an ensemble.

The two scenarios predict clearly different outcomes. The scenario that aims at suppression has both smaller mortality (670 vs 5300 deaths in a year) and lower restriction day index (72 vs 111 RDI) than the one aiming at mitigation. Its health care intensity is also smaller, as there is less need to build new intensive care units (300 vs 1050).

In addition, three other scenarios were calculated. One looked at permanent implementation of the current strict mobility restrictions (50%) and predicted a suppression of the epidemic before the end of 2020 (Figure S-1, also shown in Figure 4). Two other scenarios looked at mild restrictions from the start (25%) or relaxation of restrictions before summer (to 30%); both predicted a rapidly increasing epidemic (Figure S-2 and S-3, respectively).

**Figure 4.**
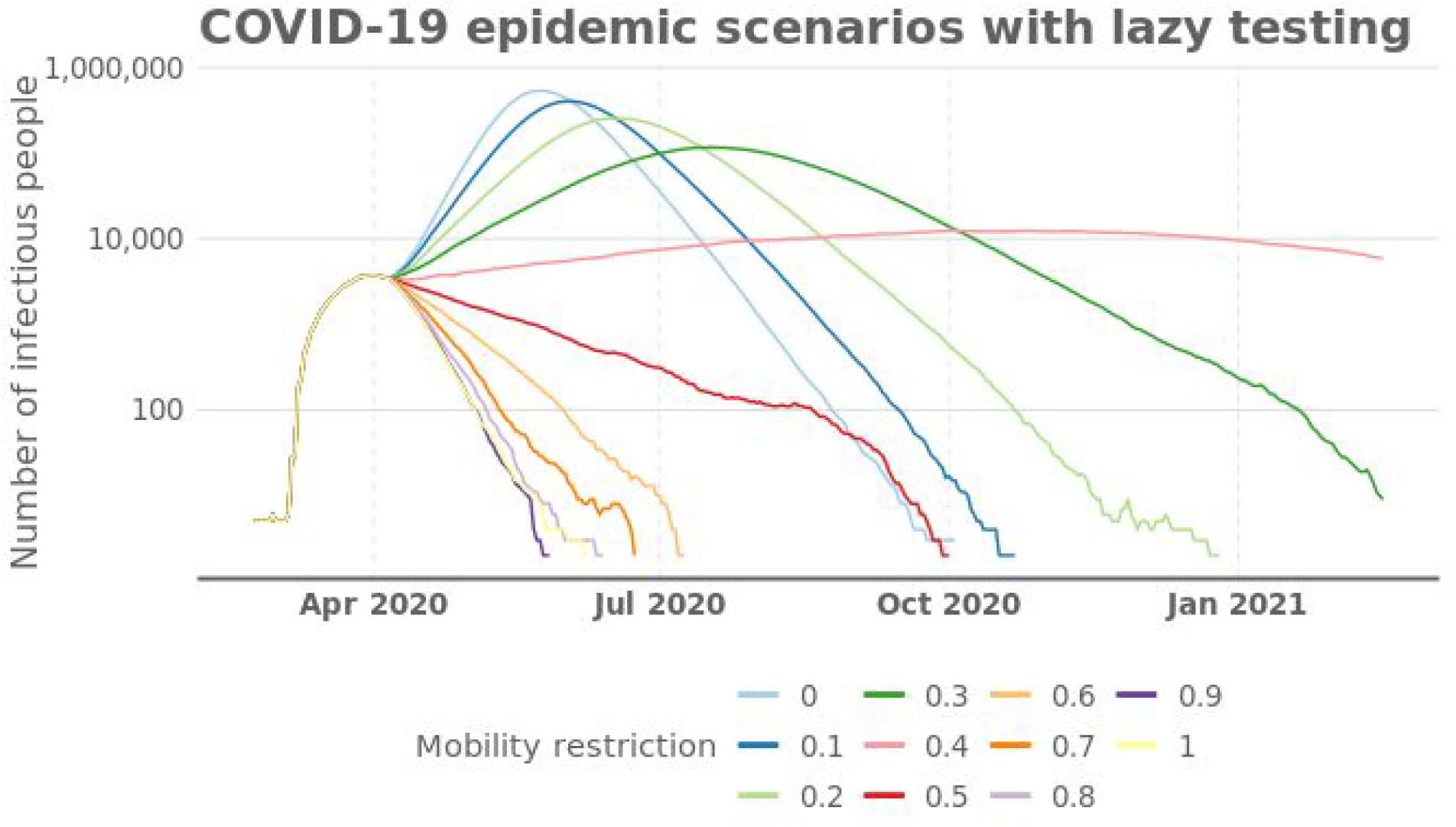
Number of active (infective) COVID-19 patients with different mobility restrictions and lazy testing (only severe cases are tested).

Figure 4 shows several potential outcomes on a timeline with lazy testing policy and varying degrees of mobility restrictions. When restriction is ca. 41%, the number of infective cases stays constant. This value was used in the sensitivity analyses to introduce maximal sensitivity of the outcomes to the variables tested. Note the logarithmic scale: without restrictions, the number of patients would be hundred times larger in early June than now (early April). According to the model, the disease can be eradicated before September 2020, if mobility restrictions are 50% or more. In contrast, if restrictions are 20% or less, the disease will spread into the population and flame out by the end of 2020.

If the mitigation strategy is adjusted for health care capacity, the mobility restrictions should be ca. 35–40% (Figure 2). In such a case, the epidemic is estimated to extend far beyond the one-year time span that is modelled here. The impacts to other sectors are not estimated here, but they are likely to be severe and get worse when the restrictions are prolonged.

Mobility restrictions may have a large impact on the number of COVID-19 patients: before February, 2021, the epidemic may infect as much as 70% or as low as 0.5% of the population, if the mobility restrictions are weak (10%) or very strict (60%), respectively.

Figure 5 shows that the *combination of contact tracing* with comprehensive testing produces very significant improvement in controlling the epidemic. This improvement is particularly noticeable under mild-to-moderate general mobility restrictions. This suggests that an effective use of testing *and tracing* makes it possible to manage the epidemic with significantly lower indiscriminate restrictions and therefore with much less economic and social damage. For example, the same mortality of 1000 people results from approx. 20 percentage points lower level of general mobility restrictions.

**Figure 5.**
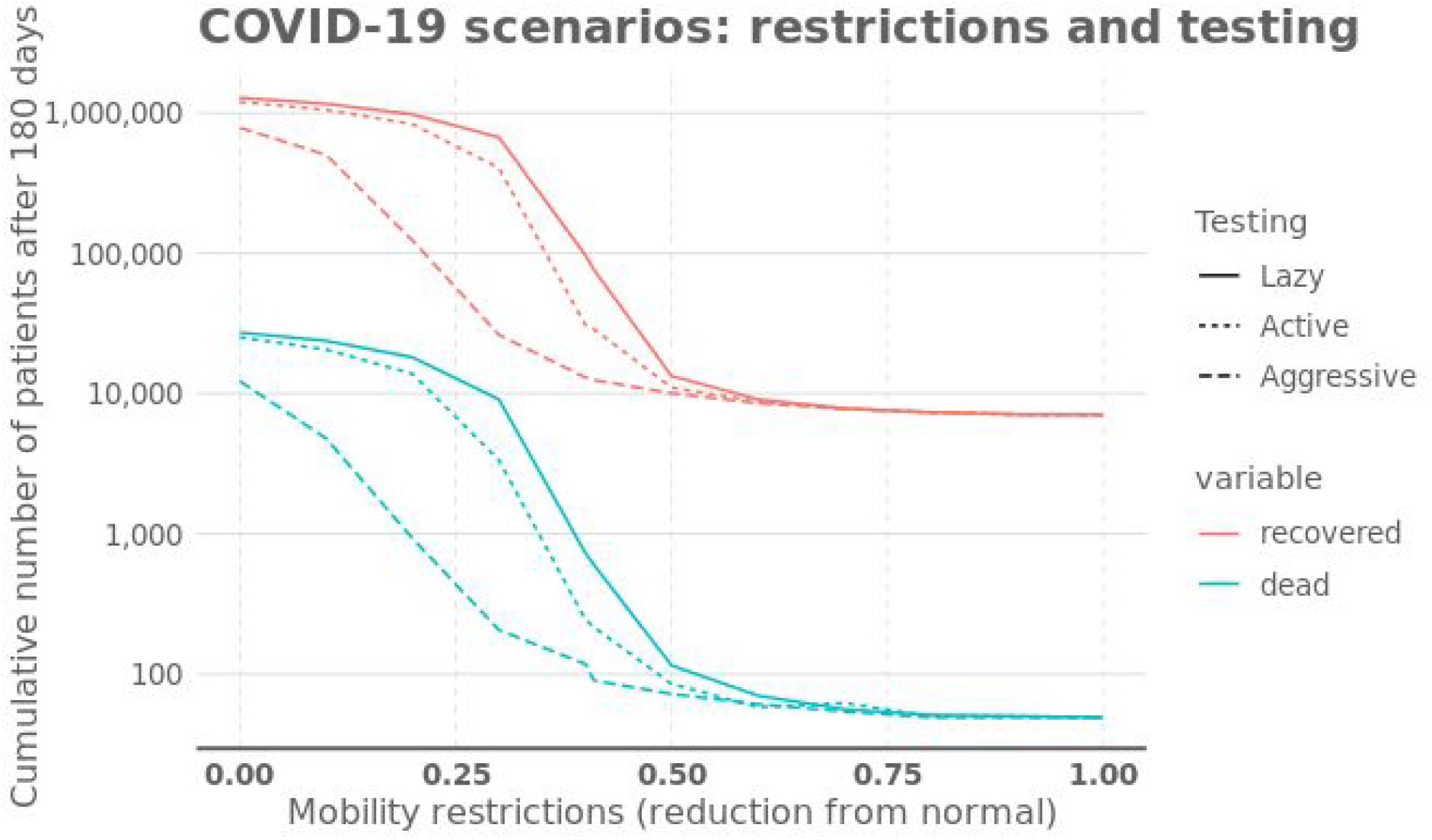
Number of COVID-19 patients after one-year simulation with permanent mobility restrictions that are adjusted on April 6th, 2020. One of the three testing policies are implemented in addition to the mobility restrictions. The accuracy parameter of Aggressive testing used for the graph is 50%.

Aggressive testing would clearly reduce the number of deaths and recovered patients (Figure 6). The death toll would be 1085, 267, and 90 with lazy, active, and aggressive testing policies, respectively, by the end of February, 2021. The number of recovered patients would be 130000, 35000, and 12000, respectively. Active testing also includes mild cases, and aggressive testing also traces all contacts of positive cases. These are optimistic numbers, as the mobility restriction in Figure 6 is strict (41% permanently), which in practice prevents the expansion of the epidemic. Lazy testing was until recently the current policy in Finland with only severe cases tested, but due to increased capacity, it has been active since early April, 2020.

**Figure 6.**
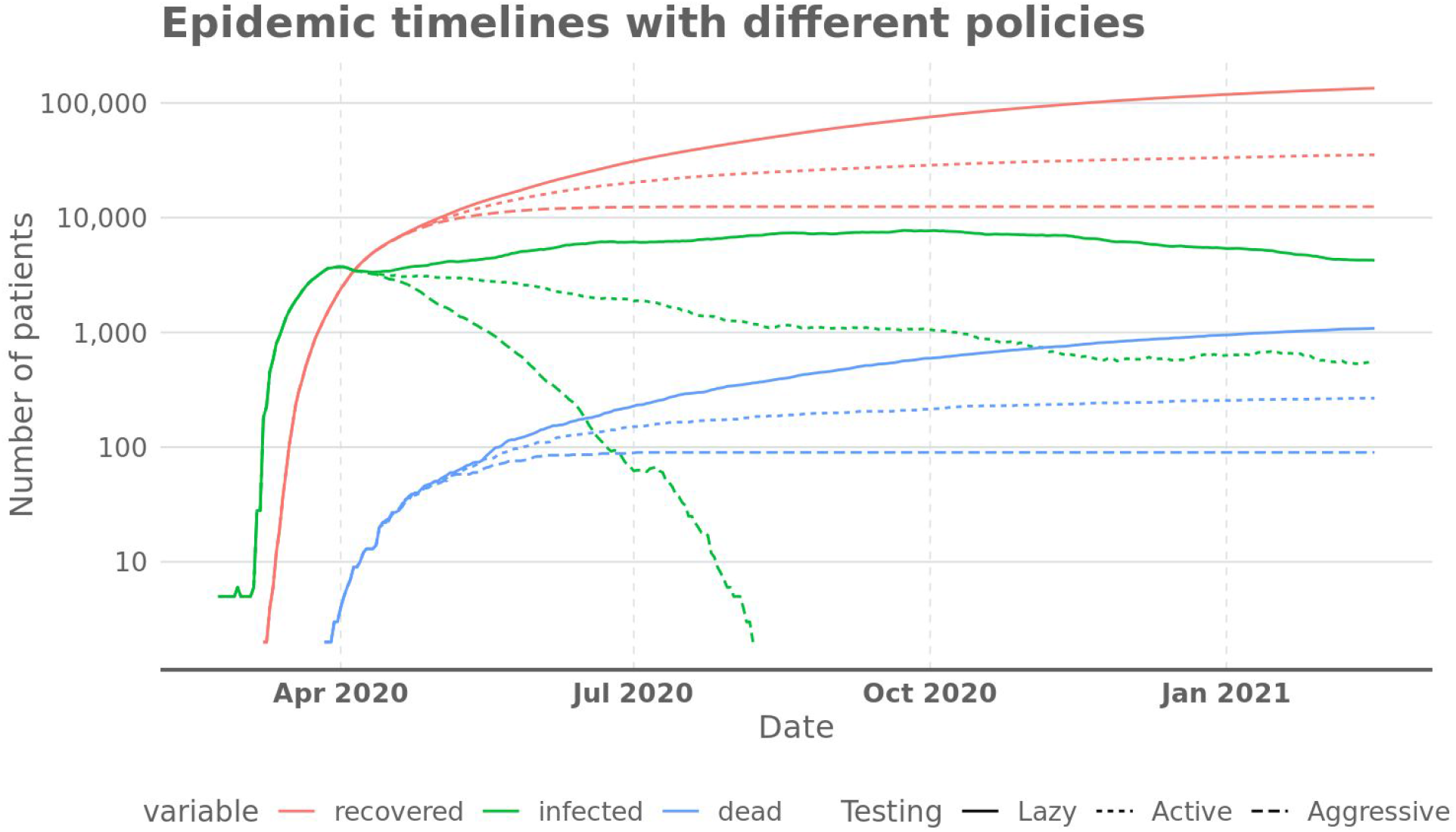
Number of active, infectious cases, recovered, and COVID-19 deaths as a function of time, with 41% mobility restriction (resulting in 137 RDI) and one of the testing policies applied permanently since April 6th, 2020.

The mitigation scenario that has 41% mobility restriction does not eradicate the disease from the population. However, a strong suppression strategy with aggressive testing can reduce the number of active cases down to hundreds within three months. In such a situation, everyday life can be more or less back to normal except for constant search for disease chains and asymptomatic patients among the population.

In particular, it is again seen that the *realising the true benefits of wider testing requires implementing also contact tracing*. Indeed, wider testing alone reduces the number of deaths by approx. 75 percent, while adding contact tracing to the mix brings further reduction of approx. 92 percent.

Another mitigation scenario allows a larger epidemic (Figure 7) to keep the restriction days lower (111 RDI) than in Figure 6 (137 RDI). Also, a suppression scenario in Figure 8 is clearly less strict with mobility (72 RDI) but is able to keep the epidemic under control (600–700 deaths in a year).

**Figure 7.**
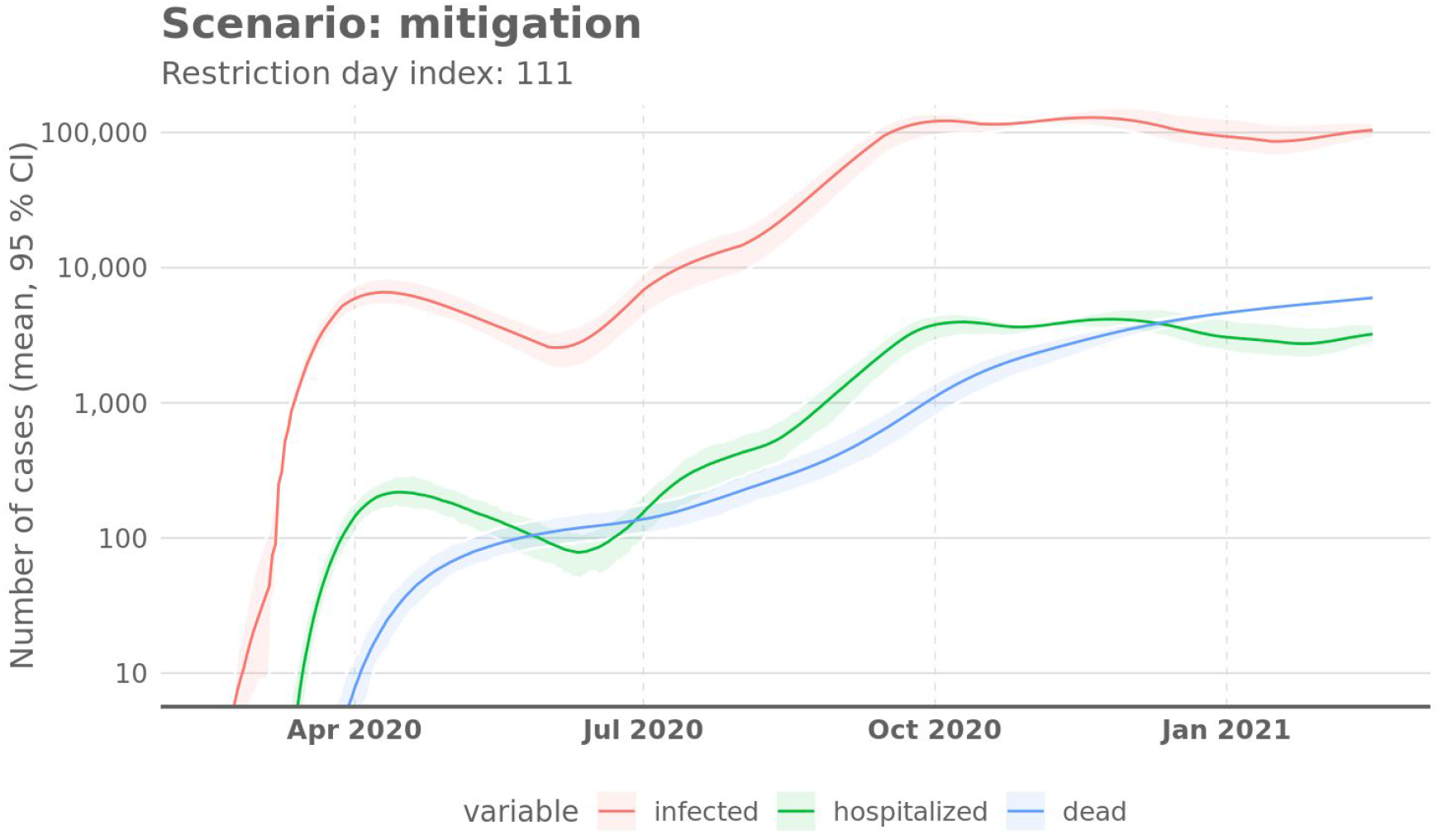
Numbers of cases in a scenario under the mitigation strategy. It contains aggressive building of hospital capacity and mobility restrictions (varying between 30–40%) just enough to prevent shortage in the capacity (a common suggestion in Finnish discussion in mid-March, 2020). Mean and 95% confidence intervals of the random variation produced by the modeling approach. They do not incorporate parameter uncertainties.

**Figure 8.**
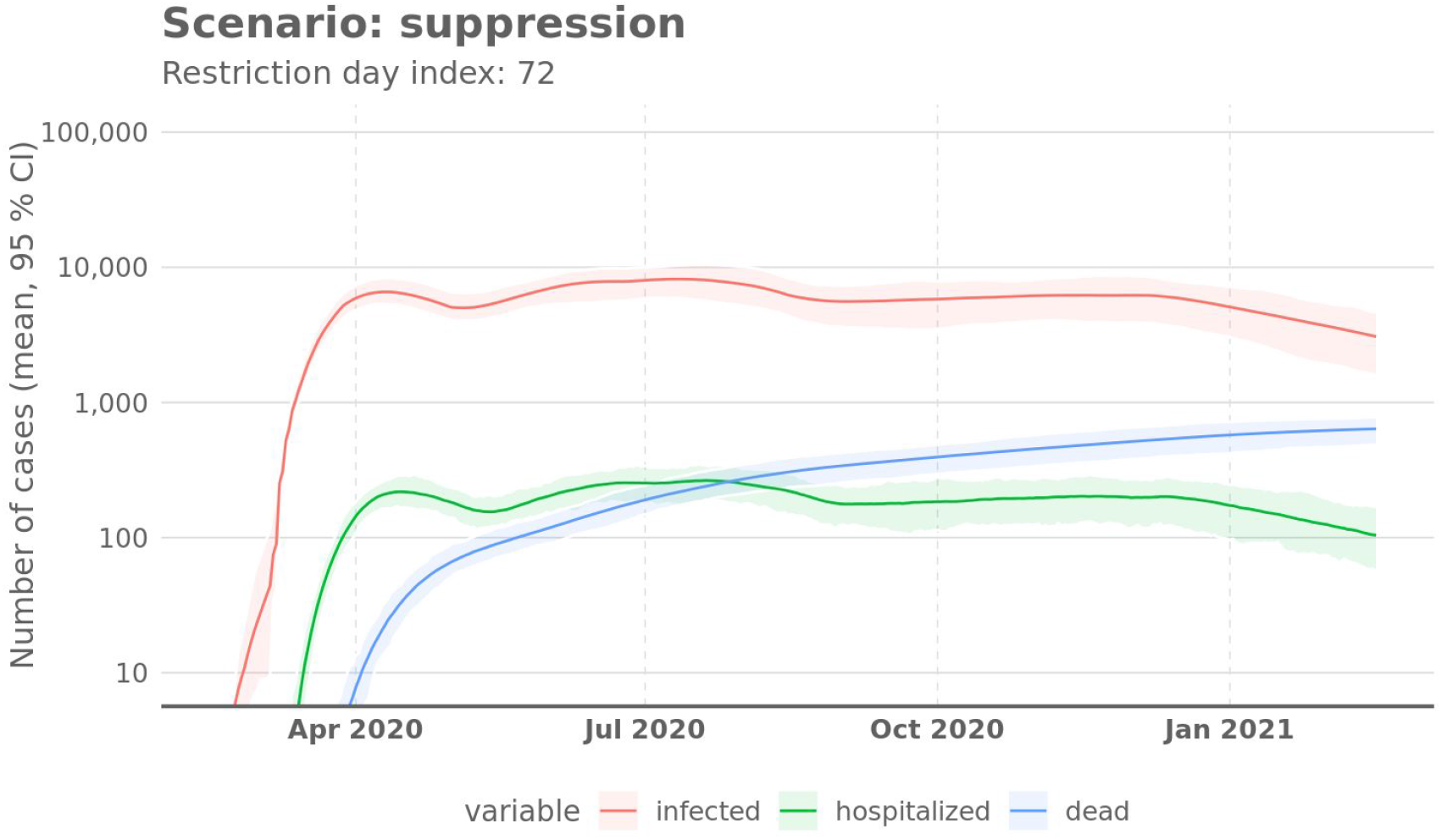
Numbers of cases in a scenario under suppression strategy. It contains initially 50% mobility restriction (the situation in Finland on 2020-04-01) but then loosening restrictions to 20–25% as aggressive testing and tracing becomes effective. Mean and 95% confidence intervals of the random variation by the modeling approach. They do not incorporate parameter uncertainties.

It is seen that the suppression strategy has lower mortality regardless of the fraction of asymptomatic cases. The difference is furthermore quite large unless at least 90 percent of cases are asymptomatic – a fraction too large to receive support from currently available data (Ferguson, 2020; ECDC, 2020).

The sensitivity analyses show that the suppression strategy is superior to the mitigation strategy at a wide range of values of asymptomatic fraction (Figure 8) and infectivity (Figure 9). It should be noted that the values far from the current best guess are very unlikely because they cannot explain the current global data about the COVID-19 epidemic.

**Figure 9.**
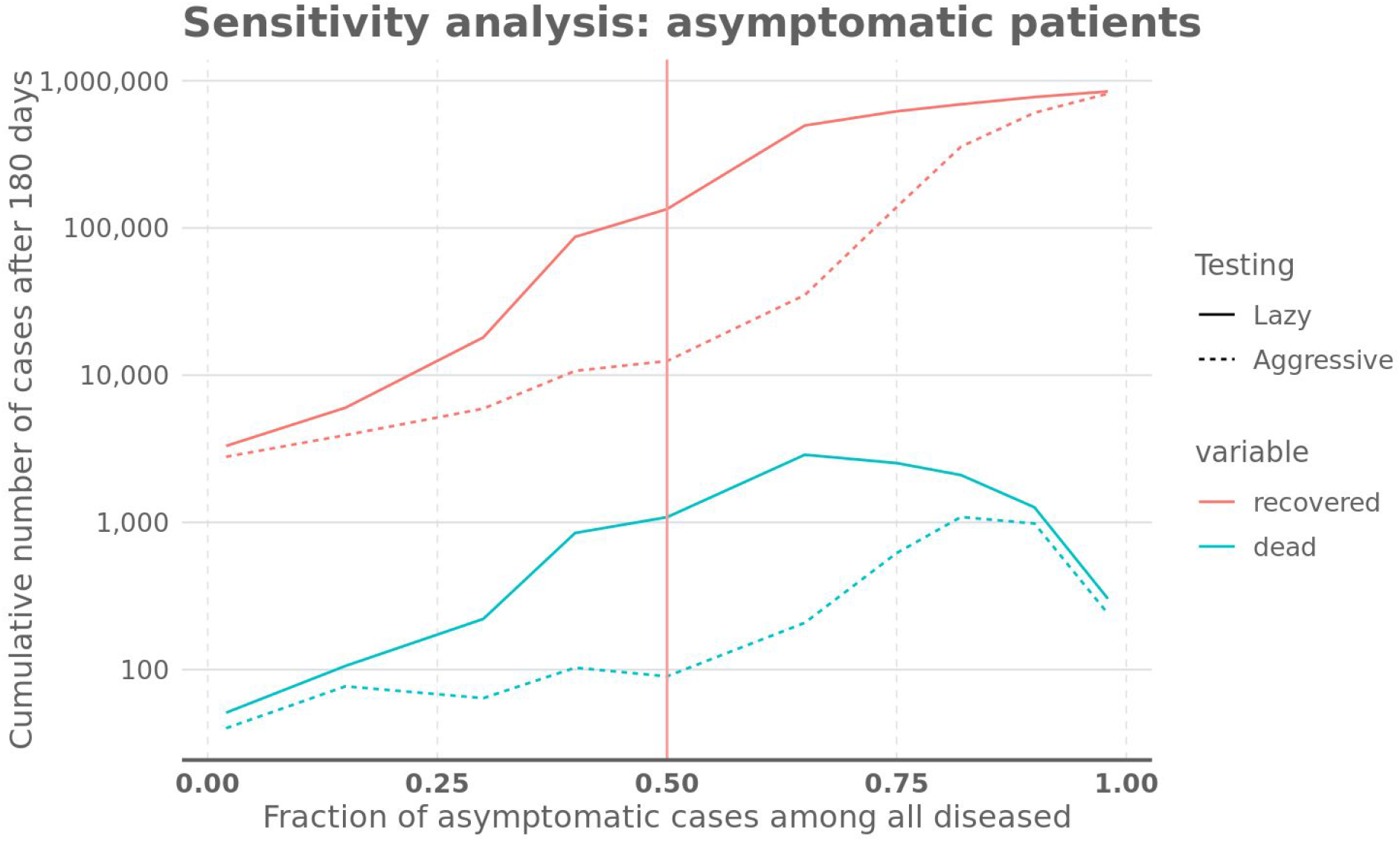
A sensitivity analysis about the fraction of asymptomatic cases with 41% mobility restriction and either lazy (in line with mitigation) or aggressive testing and tracing (in line with suppression). The values shown are after a one-year simulation. Vertical line shows the estimate used as default in the model.

## Discussion

The REINA model predicts that the suppression strategy results in a lower number of fatalities and severe cases than the mitigation strategy. Aggressive combined testing and tracing of contacts is superior to lazy testing for a wide range of mobility restrictions and also in sensitivity analyses. The difference may be tenfold or even higher. The suppression strategy has been successfully implemented in practice in China (with a delay), South Korea, Singapore, and Taiwan, thus weakening the argument that it would be infeasible. Suppression has been seen better than mitigation also in previous studies (Maharaj, 2012).

Testing makes targeted isolation of patients possible. This is important as general restrictions are detrimental for society and economy, especially if they last for several months or more. Also, the longer the restrictions, the harder it becomes to maintain them.

Suppression may consist of strong initial restrictions to get the epidemic under control (“hammer’). However, the idea is to reduce the cases enough so that active testing and contact tracing becomes feasible. The long-term objective is to prevent the spreading of the virus with testing and targeted isolation rather than general restrictions. Thus the general mobility restrictions can be reduced sooner than when the testing policy is lazy. Therefore, society gets to a more normal situation fairly soon. This diminishes the side effects of the coronavirus policies in addition to being more effective in preventing mortality (Figure 8).

In contrast, the mitigation policy allows the disease to spread in the society. The objective of the mitigation strategy is to slow down the epidemic rather than suppress it. The restrictions needed depend on the treatment capacity and how much the epidemic has to be slowed down to maintain the capacity. Unfortunately, both our modelling suggests and practical experiences from Italy, Spain, New York and elsewhere demonstrate that the capacity is very easily overwhelmed even if active restrictions are in place (Figure 7). It is noteworthy that the mitigation strategy will lead to large health impacts irrespective of actions taken, because the strategy eventually allows the disease to spread widely, and a fraction of patients will have severe outcomes. Also, restrictions are needed for a long time until herd immunity slows down the epidemic, and side effects on daily life and the economy become large. Finally, our modelling of the mitigation strategy suggests that managing the appropriate level of restrictions would be very challenging due to the slow feedback and the potential for explosive exponential growth that characterise the COVID-19 epidemic.

The probability of survival depends on the availability of hospital and intensive care. We assumed that mortality is 100% among those that need intensive care but do not get it. This assumption increases mortality when the healthcare capacity is overwhelmed. This assumption does not affect the estimates about infected people, but it critically affects the estimates about mortality.

The agent-based REINA model has some preferable properties. An arbitrary number of actions or case injections can be added on the timeline. This makes it possible to study actual situations. It is also straightforward to update parameters or adjust the model for a new population, because many parameter values are available or can be measured from a new target population, such as population age structure or an age-specific contact matrix. Also the model can be improved more easily when the parameters describe some tangible properties of the target population.

REINA does not need an estimate for R_0_. Instead, R_t_, i.e. the average number of people infected by a patient at the end of illness (time point t), is an emergent outcome of the model. The model uses explicit estimates for the three quantifiable processes that are usually embedded in R_0_, namely contacts, exposure, and infectivity.

A Finnish *contact matrix* giving empirical contact frequencies for individuals by age is available with age-specific numbers of physical contacts per day. This makes it possible to model realistic patterns of connections. They also can and have been studied in several countries, and thus country-specific models can be more realistic than relying on just global R_0_ estimates. For example, a 20-24-year old Finn has on average 9.5 total contacts per day, while an Italian of the same age has 19.3 contacts per day (Mossong et al., 2008).

The excretion of the virus from a sick individual depends heavily on the state of the disease, and unfortunately COVID-19 spreads already before a patient has symptoms and is aware of the need to avoid contacts (He et al., 2020). Aggressive testing of the contacts of the infected individuals improves the possibility of isolating also still asymptomatic cases. This is a complex interaction where some actions such as hygiene instructions can affect this parameter and thus R in a way that can be estimated with agent-based models more easily than with traditional SEIR models.

The third parameter affecting R is the probability of infection given exposure to the virus. Currently there is no published data about the value of this parameter. However, it is possible to indirectly estimate and adjust this number by comparing the model prediction with actual values of infections, hospitalizations, and deaths in the target population. Emerging data is likely to limit the plausible range of this variable in the near future.

Sensitivity analyses were performed for three key parameters, namely mobility restrictions that can be implemented (Figure 6), fraction of asymptomatic patients (Figure 9) and the probability of infection given virus contact (Figure 10). These parameters had drastic effects on the rate of disease spread and thus model outcomes. Yet, the suppression strategy implementing aggressive testing in combination with contact tracing was never worse and often clearly better than the alternative with lazy testing. Suppression with aggressive testing also requires a shorter time with strict mobility restrictions than mitigation. This makes suppression a better choice also in terms of societal and economic impacts, although this study did not attempt to quantify them.

**Figure 10.**
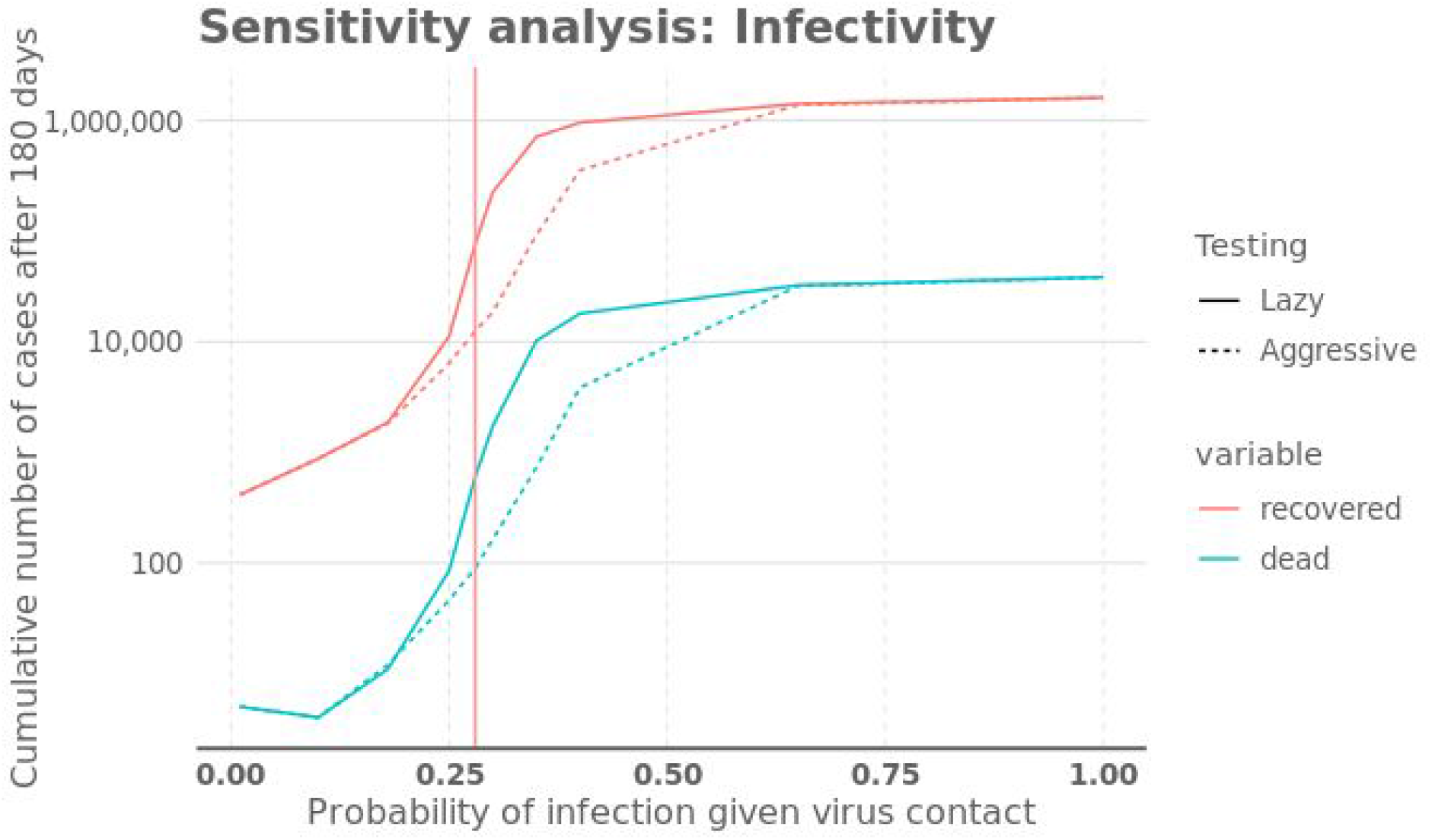
A sensitivity analysis about the probability of infection with 41% mobility restriction and either lazy (in line with mitigation) or aggressive testing and tracing (in line with suppression). The values are after a one-year simulation. Vertical line is the calibration used in the model to achieve the approx. 2.2 value for R_0_.

The preference order did not change due to scientific uncertainties tested. Although the value of information could not be estimated in this analysis because the different outcomes had no probabilities assigned to them, the sensitivity analysis implies that the value of information is actually low, i.e., the conclusion that suppression is better than mitigation is robust according to scientific evidence.

Empirical data is currently emerging about the effectiveness of actions of social isolation, closing of schools, and other mobility restrictions. The available estimates show a wide range of outcomes: Pepe et. al (2020) estimates based on mobile device location data only 19% reduction in potential encounters in Italy on 7-10 March during Italy’s third week of travel restrictions, while Jarvis et al. (2020) estimates 73% reduction of average daily contacts in the UK under lock-down in late March 2020. Google (2020) estimates 52% decrease in mobility in retail and recreation sectors in Finland on 2020-03-29 from the baseline. It is difficult to make a practical interpretation about what relative reduction is achieved by which particular action. Yet, the findings of Jarvis et al. suggests that realistic but not complete lock-down measures can indeed lead to very significant relative reduction of mobility. Bayesian and other methods could be tested in updating parameter estimates by using data from countries with different strategies.

Overall, the analysis suggests that if decision makers prefer the mitigation strategy over the suppression strategy, they have some objectives or premises that are not covered in the current agent-based model, which only looks at health outcomes.

These putative objectives are unlikely to be economic impacts of shutdowns, because both strategies are equally strict in the beginning but the suppression strategy alleviates the shutdowns earlier than the mitigation strategy.

However, some putative premises are plausible. The decision makers may ignore the health and economic benefits of suppression if they believe that testing and tracing is technically or logistically impossible or ethically unbearable due to, e.g., privacy issues. Suppression can be seen as problematic because it is required until a vaccine or cure is available, and there is no guarantee of the timing. In addition, the decision maker may believe that the disease will inevitably spread to a large proportion of the population, because success would be needed in every country before any country is safe.

A belief that the risk groups can be effectively isolated while the disease spreads to the younger population strengthens this view. However, experience from Finland and from abroad suggests that isolation of elderly population and other risk groups is unlikely to succeed, rendering such belief rather dangerous.

The good news is that there is an increasing number of practical examples where countries have shown capacity to increase both testing and tracing and where countries have effectively suppressed the disease. There are also several open-source tracing software becoming available, which will facilitate the very laborious tracing of the contacts of coronavirus positive patients.

These results make it warranted to conclude that the mitigation strategy of COVID-19 is indeed a destructive policy and should not be recommended. WHO also has systematically recommended suppression. Yet, many countries in Europe have seemed to follow the path of mitigation, until many have changed their mind since mid-March. It would be interesting to study how scientific information has entered the epidemic decision making in such countries.

All in all, we want to emphasize that the results reached are initial and include many uncertainties, some of which we were most likely not able to identify. Our aim has been to add a new modeling approach to the national Finnish context – and later internationally – that could be compared to other modeling efforts going on in parallel. Consequently, we want to promote open science by publishing the model including its source code and default parameters used to calculate the presented scenarios. Moreover, we invite other researchers and policy makers to test this approach and give valuable feedback for further development.

An obvious goal identified for further development is that of calibration and further sensitivity analysis. We assume that calibration by applying this model to data from other countries shall give valuable information. However, the outcome is always affected by national differences like societal habits and other social-distance-related parameters. On the other hand, the epidemic in Finland is just starting and therefore the time-series needed (test positives, hospitalized and deaths) is still very short. With these obstacles in mind, we invite other researchers to contribute to this important aspect by contributing their knowledge and solutions to provide the best possible calibration procedures. We have identified Bayesian approach potential for this, for example.

A particularly attractive opportunity for further development given the nature of agent-based modelling is to build increasingly realistic representations of both the population and various policy interactions. Currently the model assumes homogeneous mixing of the population contact network; in the next phase this should be improved to a more realistic model. Already existing empirical data (e.g., Mossong et al. 2008) makes it possible to model separately interactions between agents in various contexts such as workplace or education. Further work can be carried out to build a realistic household structure of population, possibly including occupational statistics. This would allow much more accurate modelling of family-level isolations and impacts of school closures. With a sufficiently detailed model of occupations the model should be able to address sector-specific restrictions such as mandating telecommuting in sectors where feasible or restaurant closures. Additionally, the agent-based approach is well adapted to still more realistic and accurate modelling of testing, contact tracing and isolation policies as well as their resource needs.

Another possible goal in the future is integration to different data sources. In order to test and use this model in different geographical areas, the local demographic and mobility (contact) data and parameters need to be imported to the model. Possible sources for this include data from Google mobility solutions, for example. Ideally, we could approach modeling where the input data for the model is updated in real time.

Fourth, the REINA model could be integrated with other models, which are able to calculate social and economic consequences of the disease as well as those of the mobility restrictions. New indicators need to be deployed, e.g. the years of life lost. In this way, the decision makers in the future would be able to balance medical, social and economic consequences in a rational yet humane way.

The model is published online for anyone to test and analyse. Also the code is openly available with an open license. The objective is to help anyone critically evaluate the details and find possible mistakes and shortcomings. Also, those who find the model useful may freely copy it for their own purposes and develop it further. In an ideal situation, an international co-creation community is formed around the model to discuss, synthesise information, and develop the model to better support the information needs of decision makers in Finland and abroad. There is an urgent need for practical policy advice in all countries about how to best tackle the COVID-19 crisis.

## Data Availability

All data and code used is available at Github.

https://github.com/kausaltech/reina-model/

https://github.com/jtuomist/corona/

## Acknowledgements

We want to thank the co-creation community and active citizens, who have worked on this topic; screened, criticised, and summarised scientific research, expert estimates, and political statements; and shared this accumulating knowledge with their fellows. Especially we thank Open Knowledge Finland and professor emeritus Matti Jantunen for organising some of this co-creation, and Tuomas Aivelo, Leo Lahti, and Juuso Pesälä for valuable comments.

## Disclaimer

The authors alone are responsible for the views expressed in this work and they do not necessarily represent the policy or views of the Finnish Institute for Health and Welfare (THL).

## Supplementary material

**Figure S-1.**
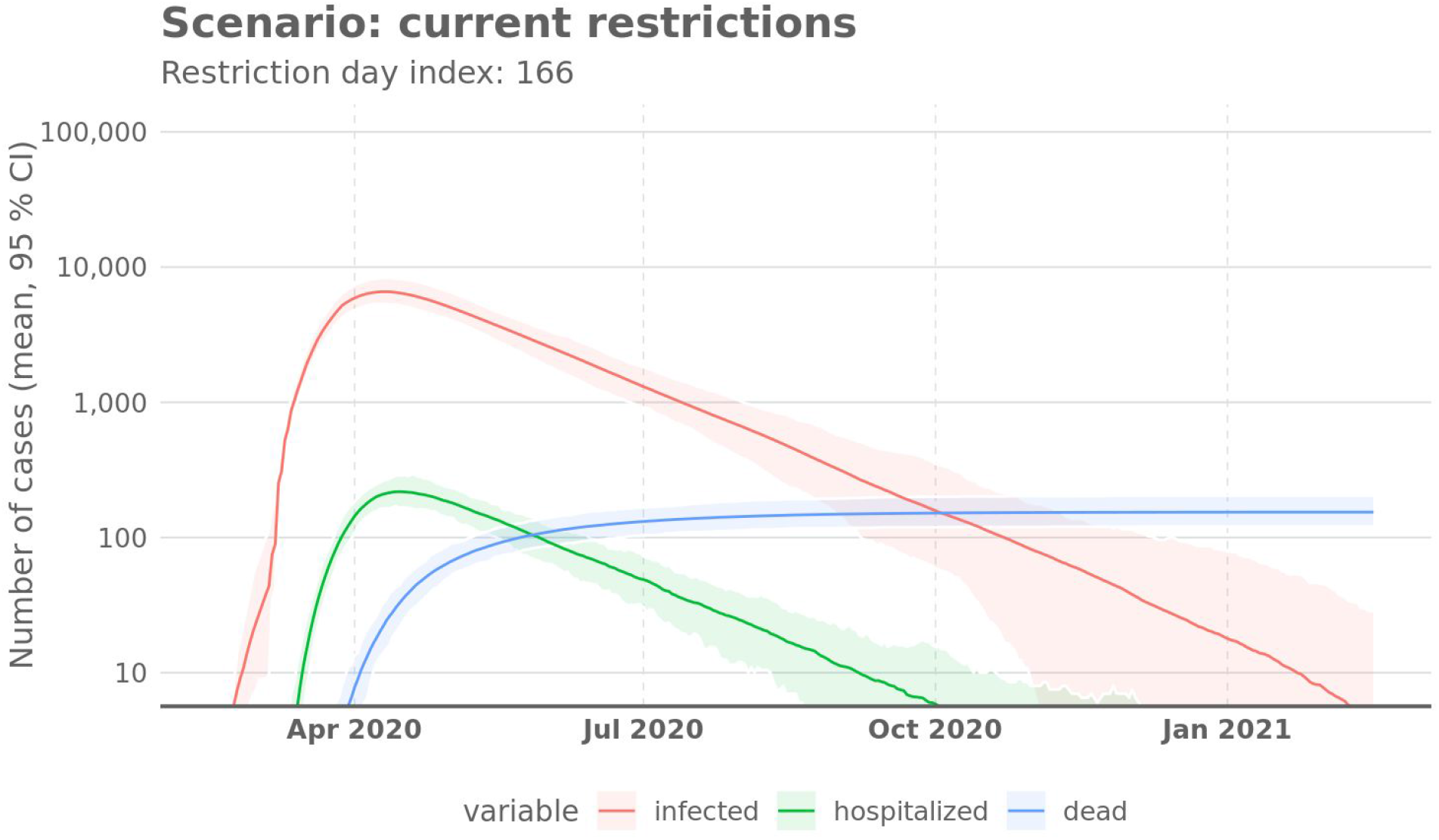
Numbers of cases of the scenario with permanent 50% mobility restriction (which is the situation in Finland on 2020-04-01), mean and 95% confidence intervals of the random variation.

**Figure S-2.**
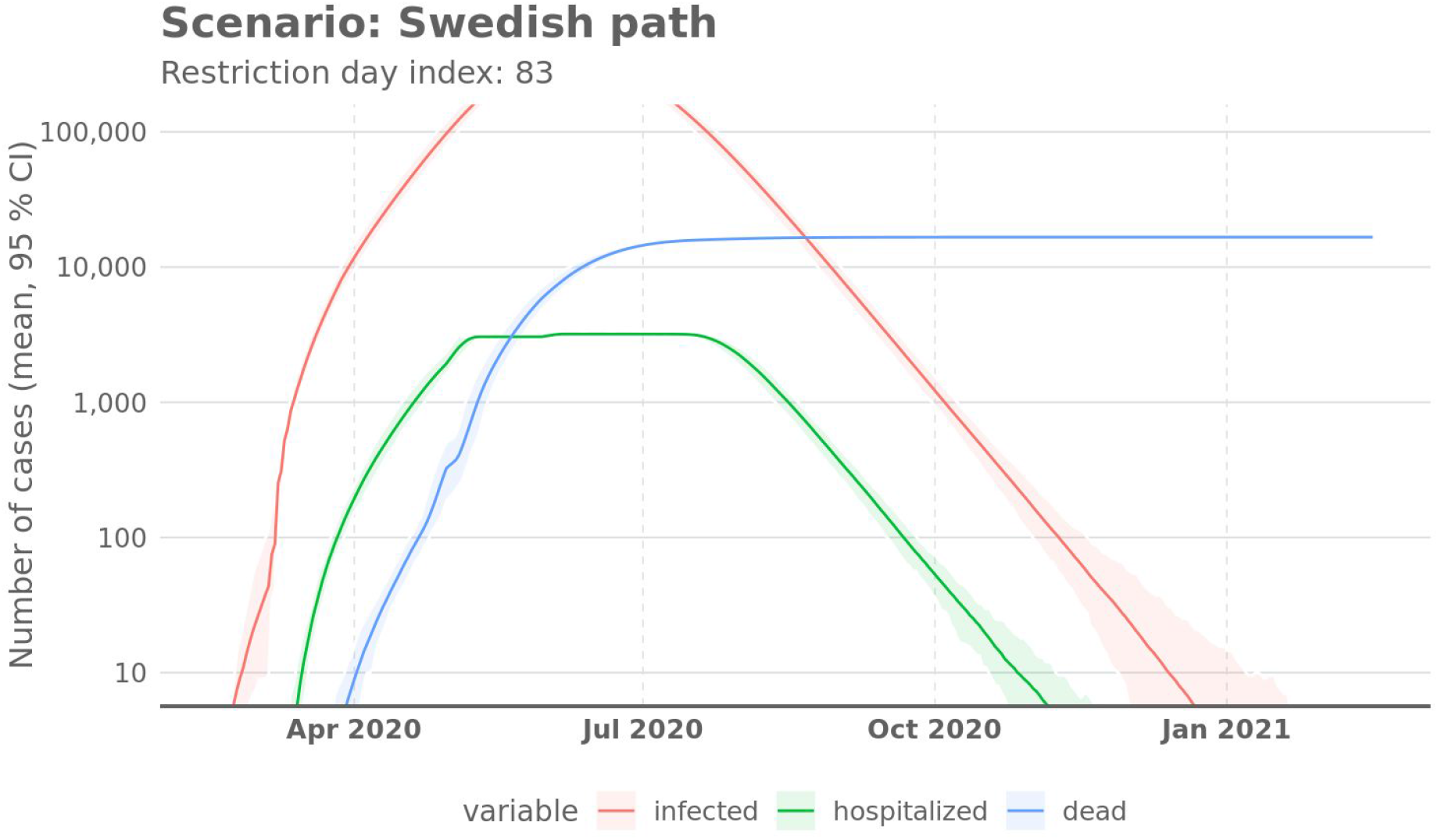
Numbers of cases of the scenario with permanent 25% mobility restriction (which was the situation in Sweden on 2020-04-01 [Google, 2020]), mean and 95% confidence intervals of the random variation.

**Figure S-3.**
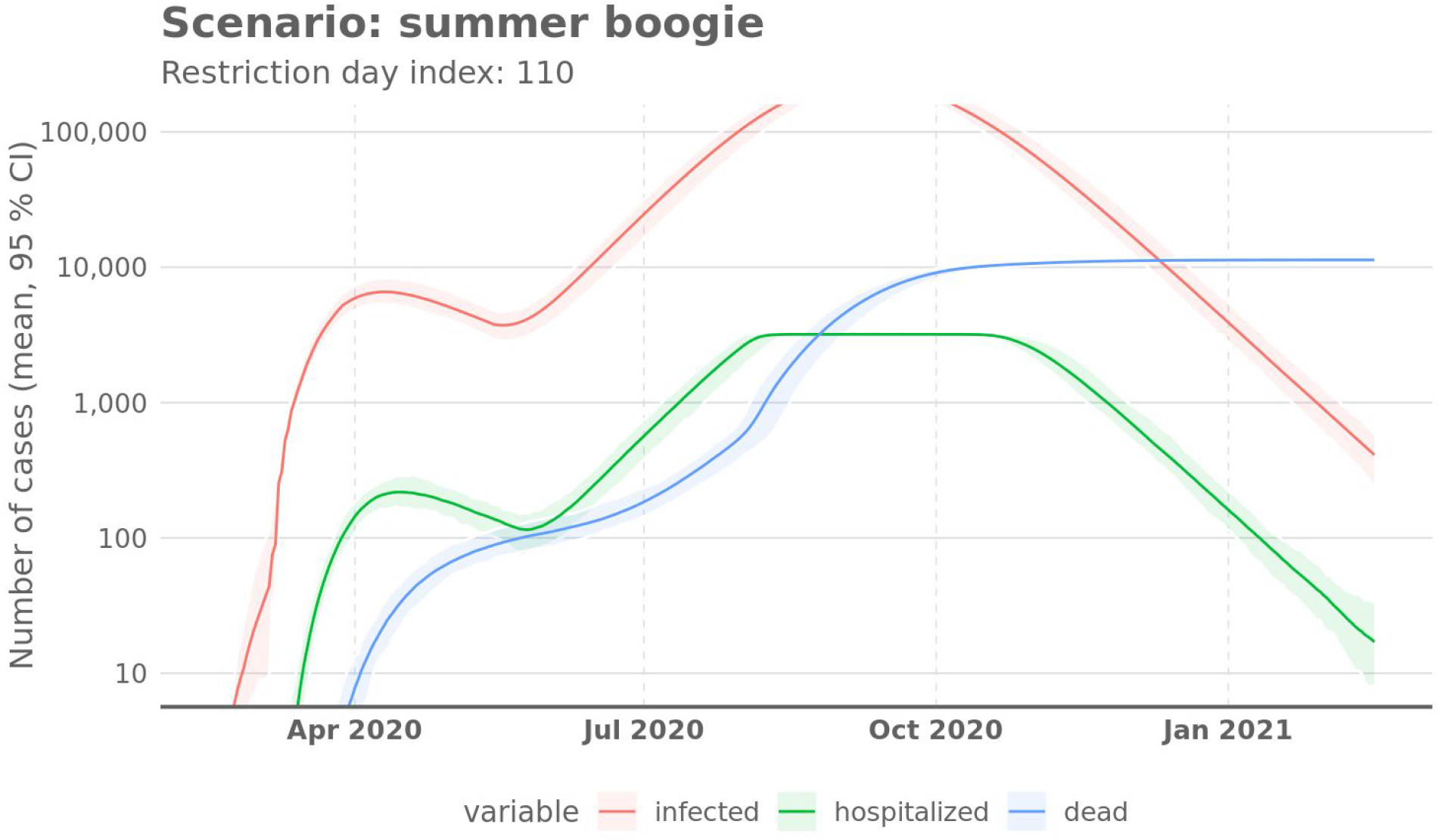
Numbers of cases of the scenario with 50% mobility restriction until 2020-05-15 and since then 30% (a suggestion heard in Finnish policy discussion). Mean and 95% confidence intervals of the random variation.

## Notes

### Competing Interest Statement

The authors have declared no competing interest.

### Funding Statement

No external funding was received.

## References

Anttila V-J. Interview in journal Ilta-Sanomat, 2020-03-30. https://www.is.fi/kotimaa/art-2000006457920.html. Accessed 2020-04-08

Boseley, S. (2020) New data, new policy: why UK’s coronavirus strategy changed. The Guardian. https://www.theguardian.com/world/2020/mar/16/new-data-new-policy-why-uks-coronavirus-strategy-has-changed Accessed 2020-04-08.

ECDC. Outbreak of novel coronavirus disease 2019 (COVID-19): increased transmission globally – fifth update, 2.3.2020.

https://www.ecdc.europa.eu/sites/default/files/documents/RRA-outbreak-novel-coronavirus-disease-2019-increase-transmission-globally-COVID-19.pdf. Accessed 2020-04-08.

Ferguson NM, Laydon D, Nedjati-Gilani G, Imai N, Ainslie K. (2020) Impact of non-pharmaceutical interventions (NPIs) to reduce COVID-19 mortality and healthcare demand. DOI: https://doi.org/10.25561/77482

Flaxman S, Mishra S, Gandy A, Unwin HJT, Coupland H et al. Report 13 - Estimating the number of infections and the impact of non-pharmaceutical interventions on COVID-19 in 11 European countries. MRC Centre for Global Infectious Disease Analysis: published online 2020-03-20. https://doi.org/10.25561/77731. Accessed 2020-04-08.

Ghenreyesus, T.A. (2020) WHO Director-General’s Opening Remarks at the media briefing on COVID-19 on 25 March 2020.

https://www.who.int/dg/speeches/detail/who-director-general-s-opening-remarks-at-the-media-briefing-on-covid-1925-march-2020. Accessed 2020-04-08.

Google. (2020) COVID-19 Community Mobility Report. Finland, March 29, 2020. https://www.gstatic.com/covid19/mobility/2020-03-29_FI_Mobility_Report_en.pdf. For all countries https://www.google.com/covid19/mobility/. Accessed 2020-04-08.

He X, Lay EHY, Wu P, Deng X, Wang J, et al. (2020) Temporal dynamics in viral shedding and transmissibility of COVID-19. MedRXiv. https://doi.org/10.1101/2020.03.15.20036707. Accessed 2020-04-08

Italian Ministry of Health. (2020) Report of the Italian Ministry of Health: Aggiornamento 08/04/2020.

https://github.com/pcm-dpc/COVID-19/blob/master/schede-riepilogative/regioni/dpc-covid19-ita-scheda-regioni-20200408.pdf Accessed 2020-04-08.

Jarvis, C.I. (2020) Impact of physical distance measures on transmission in the UK. https://cmmid.github.io/topics/covid19/current-patterns-transmission/comix-impact-of-physical-distance-measures-on-transmission-in-the-UK.html Accessed 2020-04-08.

Maharaj S, Kleczkowski, A. (2012) Controlling epidemic spread by social distancing: Do it well or not at all. BMC Public Health 12, 679. https://doi.org/10.1186/1471-2458-12-679

Mossong J, Hens N, Jit M,Beutels P, Auranen K, et al. (2008) Social contacts and mixing patterns relevant to the spread of infectious diseases. PLoS Med 5(3) e74. doi:10.1371/journal.pmed.0050074

Normile D. Coronavirus cases have dropped sharply in South Korea. What’s the secret to its success? Science Mar. 17, 2020. doi:10.1126/science.abb7566

Pepe, E. (2020) COVID-19 outbreak response: first assessment of mobility changes in Italy following lockdown (First report). https://covid19mm.github.io/in-progress/2020/03/13/first-report-assessment.html Accessed 2020-04-08.

Pueyo T. Coronavirus: The Hammer and the Dance. What the Next 18 Months Can Look Like, if Leaders Buy Us Time. https://medium.com/@tomaspueyo/coronavirus-the-hammer-and-the-dance-be9337092b56 Accessed 2020-04-08.

Tuomisto JT, Pohjola MV, and Rintala TJ. (2020) From open assessment to shared understanding: practical experiences. Health Research Policy and Systems 18: 36. https://doi.org/10.1186/s12961-020-00547-3

Verity R, Okell LC, Dorigatti I, Winskill P, Whittaker C et al. Estimates of the severity of COVID-19 disease. MedRXiv, posted 2020-03-13. https://doi.org/10.1101/2020.03.09.20033357. Accessed 2020-04-08.

Walters CE, Meslé MMi, Halla IM. (2018) Modelling the global spread of diseases: A review of current practice and capability. Epidemics 25: 1–8. https://doi.org/10.1016/j.epidem.2018.05.007

Wilson N, Barnard LT, Kvalsvig A, Verrall A, Baker M. (2020) Modelling the Potential Health Impact of the COVID-19 Pandemic on a Hypothetical European Country. MedRXiv, posted 2020-04-08 https://doi.org/10.1101/2020.03.20.20039776

